# Exploring the Challenges, Barriers and Opportunities for Neglected Tropical Disease Programmes of Conflict-affected Countries in sub-Saharan Africa: A Qualitative Investigation

**DOI:** 10.1101/2025.08.15.25333777

**Authors:** Emma M Harding-Esch, Francesca Bird, Ambreen Bashir, Louise A. Kelly-Hope

**Affiliations:** Clinical Research Department, London School Hygiene and Tropical Medicine. London, UK; Institute of Infection, Veterinary and Ecological Sciences, University of Liverpool, UK

**Keywords:** . Neglected tropical diseases, conflict, violence, sub-Saharan Africa

## Abstract

**Background:** Insecurity and conflict threaten achievement of global Neglected Tropical Disease (NTD) targets. Identifying conflict-associated challenges, and strategies to mitigate them, can inform NTD programmatic decision-making.

**Methods:** In-depth interviews were conducted with key stakeholders, including NTD programme managers and implementing partners, to explore their experiences of implementing NTD programmes in conflict-affected regions in sub-Saharan Africa. Study 1 focused on Nigeria and Study 2 on multiple countries. Thematic analysis was used to identify and extract key themes.

**Results:** Fourteen participants (7 participants per study) from six countries and several NTD programmes reported an array of complex challenges, which were categorised into three common programme-specific activity themes (implementation, community engagement, sustainability) and five external influencing themes (access to areas, water and sanitation, hygiene, inclusivity, stakeholder partnering, financial resources). Several sub-themes (e.g. surveillance, community education), unique challenges (e.g., displaced populations, access for women) and potential solutions (e.g., innovation, collaboration) were reported.

**Conclusion:** There are many complex interacting factors that hinder NTD programmes in conflict-affected NTD-endemic areas. The scale and breadth of the common and unique challenges and potential solutions require more investigation to enable the development of a practical framework to support programmes in the efforts to meet global NTD control, elimination and eradication goals.

## Introduction

Neglected tropical diseases (NTDs) are a group of 21 diseases and conditions that are an ongoing source of debilitating health, and social and economic burden to some of the most marginalised and vulnerable communities worldwide. The prevalence, distribution, and disease burden of NTDs are high in sub-Saharan Africa, accounting for more than one-third of the global burden [1]. In the last decade, NTDs have become a global health priority and the NTD Road Map 2021-2030, published by the World Health Organization (WHO), defines the global targets to control, eliminate or eradicate all NTDs, which are aligned with the sustainable development goals [2,3].

WHO has classified two broader groups for the majority of NTDs, based on the recommended method of management [1,2]. Case-management NTDs (buruli ulcer, human African trypanosomiasis, rabies, guinea worm disease, lymphoedema, leprosy, trichiasis, leishmaniasis, yaws, dengue and mycetoma), are diseases that require individual-level care due to their management complexity or lack of effective diagnostic and treatment tools. Preventative chemotherapy NTDs (onchocerciasis, schistosomiasis, lymphatic filariasis, trachoma and soil transmitted helminths), can be treated through mass drug administration (MDA) to entire populations, aiming to reduce morbidity and prevent further transmission of these diseases. Preventative chemotherapy is also implemented alongside vector control, water, sanitation, and hygiene (WASH) services and veterinary public health services, to support and improve treatment outcomes. The NTDs that do not fall into these two categories, such as rabies, snakebite and dengue, have different approaches to prevention and control.

The long-term nature of tackling many NTD infections and diseases is further compromised by limited financial and human resources and access to healthcare services in many low-income countries, especially in Africa [4]. In addition, NTD programmes are hampered by a rise in conflict and political instability that disrupt societal structures and healthcare services, resulting in: strains on the health infrastructure, including damaged buildings and disruption to essential facilities such as WASH; unintended mass population movement, including refugees and internally displaced persons living with unsanitary and crowded conditions and vulnerable to opportunistic infections; and a diversion of essential funding [5–9]. These challenging factors can alter the epidemiology of NTDs and negatively impact the effectiveness of programme activities, including conducting prevalence surveys and other surveillance activities, and implementing interventions such as preventive chemotherapy and patient clinical care, as well as WASH-related activities [10–12].

Currently, there are no formal recommendations for NTD programmes operating in conflict settings, however, there is increasing recognition among stakeholders that it is a critical challenge that needs addressing to ensure that ‘no one is left behind’ and the NTD Road Map goals are achieved and aligned with the United Nations Sustainable Development Goals [2,11–16]. There has been little research to examine how conflict events such as civil war, terrorism or military coups directly affect NTD programmes, and how programmes have responded to these challenges. Therefore, this study aimed to engagement with those involved with NTD programme management and implementation activities as an important first step to determine the range of challenges and potential mitigating strategies.

## Methods

### Study overview

Two separate, but complementary, qualitative studies in conflict-affected countries in Africa were conducted in July-August 2023. Study 1 focused on one county, Nigeria, to gain an in-depth exploration within a single setting. A variety of key stakeholders, defined as individuals working for organisations supporting NTD programmes in Nigeria, participated in the study. . Study 2 focused on multiple countries and involved one NTD programme manager in each country. This enabled cross-country diversity and commonality of perspectives and experiences to be identified.

### Participant recruitment

A proposed sample size of 10-15 participants was targeted for each study, or when saturation was reached. For Study 1, key stakeholders working on NTD programmes in Nigeria were recruited. For Study 2, the NTD programme manager from 16 African countries that were classified as high conflict and/or high NTD burden countries [11,12] were invited to participate. The countries invited to Study 2 participation included Angola, Burkina Faso, Burundi, Cameroon, Central African Republic (CAR), Chad, Democratic Republic of the Congo (DRC), Ethiopia, Mali, Mozambique, Niger, Nigeria, Kenya, South Sudan, Tanzania, and Uganda. Participants were identified through purposive sampling based on their professional roles [17], and snowball sampling [18].

### Data collection

Participants were invited by email to participate in semi-structured interviews, using pre-determined interview topic guides (Supplementary Information 1). This design allowed participants to reveal information that may have not been considered by the researcher prior to the interview [19]. The topic guides were developed from an initial review of the literature on NTD programmes in conflict-affected countries, with a set of themes and open-ended questions established to guide the interview. The topic guide was further refined through feedback from the NTD NGO Network (NNN) Conflict and Humanitarian Emergencies (C&HE) working group [20], and the International Coalition for Trachoma Control (ICTC) Special Populations task team [16].

Interviews were conducted and recorded using online video conferencing (Microsoft Teams for Study 1, Zoom for Study 2), and lasted between thirty minutes to one hour [21]. The themes and questions outlined in the topic guide were used flexibly and adapted as interviews took place and new information was revealed by participants. In instances where participants were not proficient in English, the questions were translated into their preferred language during the interview. For French interviews, a translator was used, with the video conferencing platform recording and transcribing the interview. The transcription was cross-checked against the recording by a native speaker. For Portuguese interviews, the verbal responses were automatically transcribed by the video conferencing platform, and the recording and transcription were cross-checked. The transcription was then transcribed back into English and checked by a native speaker.

### Data analysis

Transcripts for both studies were uploaded to and analysed in NVivo (QSR International Pty Ltd. 2020). Data analysis was conducted concurrently with the interview process, so that emerging themes could inform the questions asked in forthcoming interviews [21]. A thematic analysis was performed to identify common salient experiences of NTD stakeholders, guided by Braun and Clarke’s 6-steps [22]. Codes and themes were identified both *a priori* using a deductive approach based on the research question, objectives and themes developed for the interview topic guide, and *a posteriori* using an inductive approach to recognise any unplanned or emergent themes that emerged from interviews [23]. After completion of the interviews, the transcripts were independently reviewed and analysed by a second researcher to finalise the theme and sub-theme analysis.

## Results

For Study 1, seven key stakeholders working on NTD programmes in Nigeria participated. They were mostly males and worked at the Ministry of Health or for an NGO, with a focus on trachoma For Study 2, seven NTD programme managers working for the Ministry of Health, each in a different country and working on a range of NTDs, participated, including four males and three females, with between 6 -17 years’ experience. Specific countries are referred to as Country 1-7.

### Thematic analysis

Two key categories emerged from the thematic analysis: programme-specific challenges, and external programme influences (Figure 1). Within each category, there was several themes and sub-themes.

**Figure 1.**
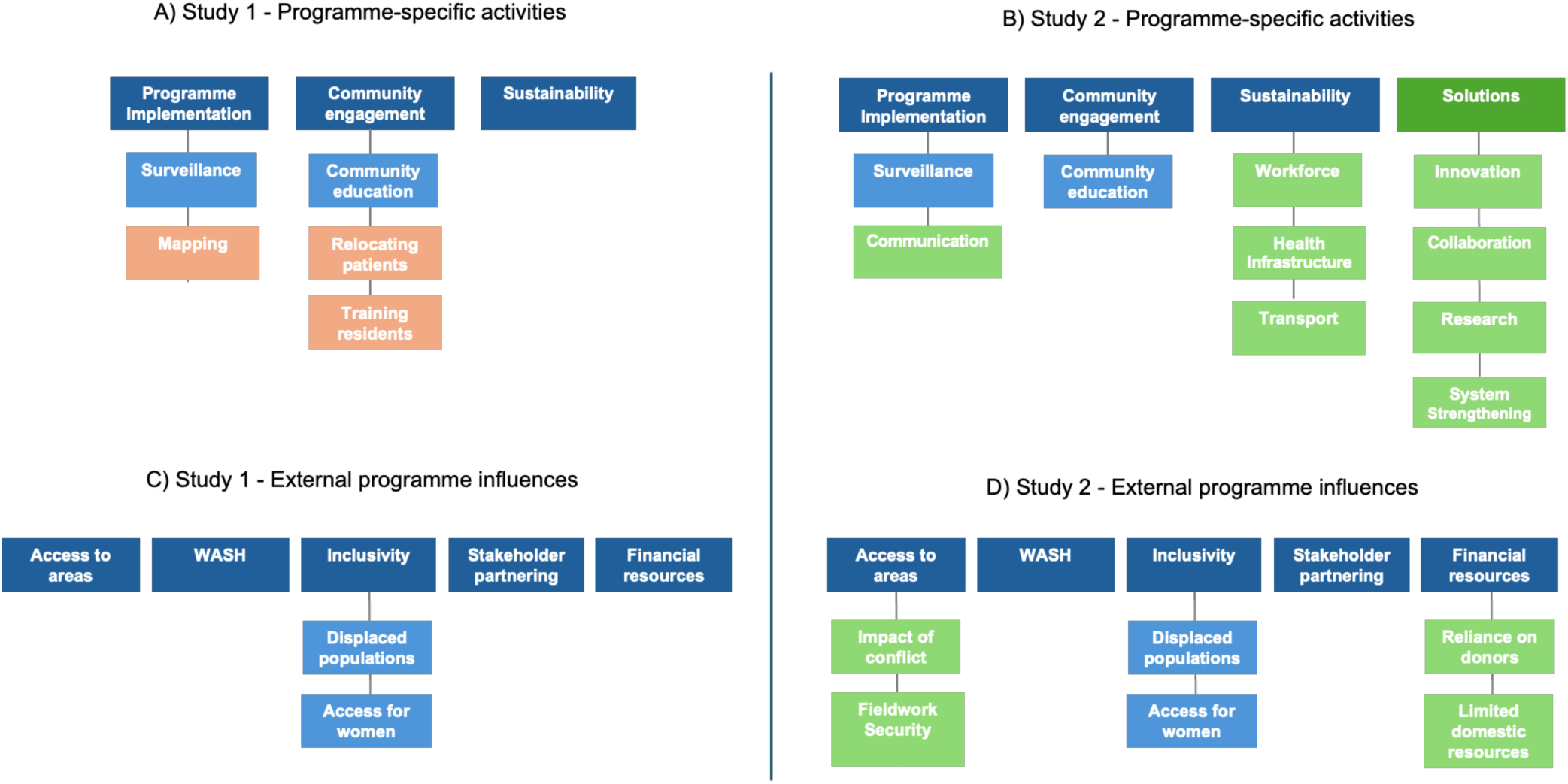
Main and sub-themes in both qualitative studies. Note: Dark blue = common themes between both studies; Light blue =common sub-themes between studies; Light orange = unique sub-theme for Study 1; Dark green = unique theme for Study 2; Light green = unique sub-themes for Study 1

### Programme-specific challenges category

There were three common programme-specific challenge themes between both studies: programme implementation, community engagement, and sustainability. Study 2 also identified potential solutions as a main theme, with four sub-themes related to innovation, collaboration, research and health system strengthening. Table 2 summarises the programme-specific challenges reported by each participant across the two studies. In addition, there were nine sub-themes: two were common between the studies (surveillance and community education); Study 1 reported mapping, relocating patients and training residents; and Study 2 reported communication, workforce, health infrastructure and transport (Figure 1 A, B). Selected key quotes are in Boxes 1-4.

**Table 1.** Summary of participants’ gender, organisation type and neglected tropical disease (NTD) focus.

| Participant | Gender | Organisation type | NTD(s) focused on in role |
| --- | --- | --- | --- |
| **Study 1. Nigeria** |  |  |  |
| 1 | Male | Ministry of Health | Trachoma |
| 2 | Male | Ministry of Health | Lymphatic filariasis, Onchocerciasis, Trachoma, Schistosomiasis, Soil transmitted helminthiasis |
| 3 | Female | NGO | Trachoma |
| 4 | Male | NGO | Trachoma |
| 5 | Male | NGO | Trachoma |
| 6 | Male | NGO | Trachoma |
| 7 | Male | NGO | Trachoma |
| **Study 2. Multiple countries** |  |  |  |
| 1 | Male | Ministry of Health | All NTDs |
| 2 | Male | Ministry of Health | Lymphatic filariasis, Onchocerciasis |
| 3 | Female | Ministry of Health | Trachoma |
| 4 | Female | Ministry of Health | Trachoma, Lymphatic filariasis, Schistosomiasis, Soil transmitted helminthiasis, Scabies, Snake bite |
| 5 | Male | Ministry of Health | Trachoma, Guinea worm, Mycetoma |
| 6 | Female | Ministry of Health | Trachoma, Lymphatic filariasis |
| 7 | Male | Ministry of Health | Trachoma |

NGO: non-governmental organisation

**Table 2.** Summary of the programme-specific challenges and external programme influences, reported by participants across the two studies.

| Participant | Programme-specific challenges |  |  | External programme influences |  |  |  |  |
| --- | --- | --- | --- | --- | --- | --- | --- | --- |
|  | Programme Implementation | Community Engagement | Sustainability | Access to insecure areas | Stakeholder partnering | Inclusivity | WASH | Funding |
| **Study 1. Nigeria** |  |  |  |  |  |  |  |  |
| 1 | X | X |  | X | X | X | X |  |
| 2 | X | X | X | X | X | X | X | X |
| 3 |  | X | X |  | X |  | X |  |
| 4 | X | X | X |  | X | X | X |  |
| 5 |  | X |  | X | X | X | X |  |
| 6 | X | X |  |  | X | X | X | X |
| 7 | X | X | X |  | X | X | X |  |
| **Study 2. Multiple countries** |  |  |  |  |  |  |  |  |
| 1 | X | X | X | X | X | X | X | X |
| 2 | X | X | X | X |  |  | X | X |
| 3 | X | X | X | X |  |  |  |  |
| 4 | X | X |  | X | X | X | X | X |
| 5 |  | X |  | X |  | X | X | X |
| 6 |  | X | X | X |  |  | X | X |
| 7 | X | X |  | X | X | X |  |  |

### Programme implementation (Common theme across the two studies)

Ten participants highlighted programme implementation as a challenge in areas of insecurity. Selected key quotes are in Box 1.

#### Surveillance (Common sub-theme)

Six of the 14 participants revealed the barriers that insecurity presents to achieving effective surveillance. Whilst participants noted that there is a great need for surveillance surveys to determine whether and what type of interventions are needed in each district, such data can be difficult to obtain in insecure areas as it is dangerous to for health personnel to go to these areas.

#### Mapping (Study 1 sub-theme)

Four participants discussed the importance of partnering with key stakeholders through baseline surveys (mapping) to optimise NTD interventions in insecure regions. Significant collaborations with the ministries of Environment, Education, and Water Resources have benefited NTD programmes by supporting activities of the National Trachoma Task Force.

#### Communication (Study 2 sub-theme/linked to health infrastructure sub-theme)

Six participants highlighted how telephone lines and internet services in conflict-affected areas may be disrupted for significant periods of time, rendering the coordination of activities and information sharing between different programme areas nearly impossible hindering implementation. This is linked to the Study 2 health infrastructure challenges sub-theme.

###### Box 1. Selected quotes on programme implementation challenges

***Surveillance***

*“Surveillance surveys to measure the effect or your results of your intervention…So in areas in districts that have security challenges, it’s very difficult to undertake these surveys.” (Study 1, Nigeria, Trachoma)*

*“It’s difficult for you to measure appropriately or accurately, what are the results of your interventions. So, we have issues in terms of getting data for service in security challenged areas.” (Study 1, Nigeria, Trachoma)*

*“Despite the challenges, the programme has seen successes through effective strategies like using mobile devices for surveillance and hit-and-run methods for surgeries” (Study 1, Nigeria, Trachoma)*

***Mapping***

*“So that’s why mapping is key. How do we do the mapping? As I said, we try to identify the key stakeholders what can we add, that can help us because of the issue of security.” (Study 1, Nigeria, Lymphatic filariasis, Onchocerciasis, Trachoma, Schistosomiasis, Soil transmitted helminthiasis)*

***Communication (infrastructure)***

*“Sometimes internet is disconnected for a year for completely affected areas, but if the economy allows us then we use satellite communication. Even phone communication is impossible in conflict affected areas and, 100% we can’t access that area, unless there is satellite communication.” (Study 2, Country 1, all NTDs)*

*“Power stations and telecommunications were vandalised, which temporarily isolated the region and disrupted communication.” (Study 1, Country 4, trachoma, lymphatic filariasis, soil transmitted disease, snake bites, schistosomiasis, scabies)*

### Community engagement (Common theme across the two studies)

All 14 participants highlighted the need for community engagement in areas of insecurity to implement successful NTD programmes. Selected key quotes are in Box 2.

#### Community education (Common sub-theme)

Five participants raised the importance of promoting community education in areas of conflict to disseminate information and improve awareness about the diseases and treatments, alongside NTD programmes. For example, educating the community about the benefits and potential side effects of the drugs and maintaining high training standards for drug distributors to ensure there are limited side effects or reactions.

#### Relocating patients (Study 1 sub-theme/potential solution)

Four participants highlighted the need to relocate patients to secure locations for effective trachoma surgeries. This process involves organising transport, conducting surgery in a safe area, and providing aftercare advice. In Nigeria, the programme funds the transport of patients from high-risk areas for surgery and post-surgery treatments.

#### Training residents (Study 1 sub-theme)

Four participants highlighted the benefit of recruiting and training local residents as community health workers to distribute drugs and collect data (using digital technologies in some cases) in areas of insecurity. Such individuals are likely to have greater access to populations in highly insecure areas.

###### Box 2. Selected quotes on community engagement challenges

***Community education***

*“Another obstacle is the lack of awareness about certain diseases, with communities and political actors often unaware of neglected diseases, attributing health issues to cultural factors. Overcoming this challenge involves leveraging community knowledge and traditional networks, using local languages to disseminate information and educate populations.” (Study 2, Country 6, Trachoma, Lymphatic filariasis)*

*“They feel that if they are with you, maybe the bandits or the whatever they may even be part of the targets.” (Study 1, Nigeria, Trachoma)*

*We have to mobilise them clear clearly on the programme, let them understand the benefit they are going to have as a result of the medication we’re going to provide to them.” (Study 1, Nigeria, Trachoma)*

***Relocating patients (potential solutions)***

*“Sometimes if the security is intense, we ask those people to come to another place. We relocate them to another place based on discussion with them.” (Study 1, Nigeria, lymphatic filariasis, onchocerciasis, trachoma, schistosomiasis soil transmitted helminthiasis.)*

***Training residents (potential solutions)***

*“We train all our staff in different security management levels, from basic security to hostile environment and defensive driving.” (Study 1, Nigeria, Trachoma)*

*“Train just the people who are residing in those areas because they understand the security situation very well. They are known to the locals, so they can easily get your data.” (Nigeria, Trachoma)*

*“Residents of insecure areas are also being trained to collect data and provide updated security information to the NTD programme officers. Training involves using electronic data collection systems such as CommCare and KoboCollect.” (Study 1, Nigeria, Trachoma)*

### Sustainability (Common theme across the two studies)

Eight participants highlighted the importance of promoting sustainable NTD programmes, particularly in areas of insecurity but provide limited further information, particularly for Study 1 where no sub-theme could be identified. Selected key quotes are in Box 3.

#### Workforce (Study 2 sub-theme)

Two participants shared their experiences of the significant difficulties that arise when the programmes are understaffed due to migration caused by conflict, and how this poses a threat to the long-term sustainability of their programme.

#### Health infrastructure (Study 2 sub-theme)

Three participants highlighted the impact of damage to health infrastructure in insecure regions, significantly impacting NTD control initiatives. Damage and inaccessibility to health facilities was reported to severely limit their ability to provide healthcare services to impacted communities.

#### Transport (Study 2 sub-theme)

Three participants reported that transport infrastructures, such as roads, bridges, are often damaged and rendered impassable due to conflict-related incidents and may hinder medical supply delivery and programme staff movement. This creates difficulties in accessing remote locations where NTDs are widespread.

###### Box 3. Selected quotes on sustainability challenges

*“Sustainability is now the issue. Because when these people are not there, I don’t know how some of the programmes continue, even though we have made a lot of progress in terms of elimination.” (Study 1, Nigeria, Trachoma)*

***Workforce***

*“Regrettably, there have been disruptions due to personnel shortages and the consequential migration of staff.” (Study 2, Country 3, Trachoma)*

***Infrastructure***

*“So, we are really suffering even if the war is stopped and it is safe to travel in those areas, the health facilities are damaged, looted, medicines are looted as they have been in the war for many years. Even after the war has stopped it’s not an easy task to resume the service because everything is damaged and distracted and one has to start from the beginning.” (Study 2, Country 1, all NTDs)*

***Transport***

*“Challenges faced during a peak time in areas heavily affected, which were inaccessible due to war.” (Study 2, Country 4, trachoma, lymphatic filariasis, soil transmitted disease, snake bites, schistosomiasis, scabies)*

### Potential solutions (Study 2-only theme)

The potential solutions for the challenges affecting NTD programmes were not widely reported, but broadly raised in relation to innovation, collaboration, research, and health system strengthening, Selected key quotes are in Box 4.

#### Innovation (Sub-theme)

Five participants reported that innovation is crucial in combating NTDs in conflict-affected areas. They emphasised the importance of integrating advanced technologies like biometric fingerprint recognition. Additionally, promoting sanitation marketing, such as local detergent manufacturing and toilet construction was reported.

#### Collaboration (Sub-theme)

Five participants reported that collaboration with non-NTD stakeholders, particularly humanitarian agencies, non-governmental organisations (e.g. UNICEF), security operators and the military was an important solution as it helped to plan and execute the programme activities, and to gain access to insecurity areas while also ensuring safety.

#### Research (Sub-theme)

Two participants mentioned that conducting research could be crucial in addressing the numerous obstacles encountered by NTD programmes in conflict settings. They emphasised that research is paramount, as it not only enhances understanding of the complex nature of various scenarios, but also provides invaluable guidance in devising effective strategies to mitigate the impact of conflicts on different aspects of society.

#### Health system strengthening (Sub-theme)

Four participants highlighted the need for health system strengthening for effectiveness and success of healthcare initiatives, particularly in areas susceptible to conflicts. They emphasised pre-emptive planning, collaborating with the education department, and tackling fundamental necessities such as WASH, to significantly strengthen systems. In turn, enhancing overall communal health, having flexibility in timelines, and being ready to adjust to evolving conditions, were considered vital.

###### Box 4. Selected quotes on potential solutions

**Health system strengthening**

*“The first thing is the system strengthening, we must strengthen the system so that we will be in a good position to cope up when conflicts happen, we could continue work in the absence of the external support.” (Study 2, Country 2, Lymphatic filariasis and onchocerciasis)*

***Collaborations***

*“… means including military personnel to facilitate their work. Collaborating with humanitarian organizations …. to gain a better understanding of the actual conditions in these areas …. This information is crucial for planning effective interventions. This collaboration with the military as an innovation, recognizing that waiting for change without taking proactive steps is not a viable option.” (Study 2, Country 4, Trachoma, Lymphatic filariasis, Schistosomiasis, Soil transmitted helminthiasis, Scabies, Snake bite)*

*“We closely collaborate with security operators to ensure thorough planning and execution.” (Study 2, Country 5, Trachoma, Guinea worm, Mycetoma)*

*“One challenge involved transportation for field teams [in conflict affect areas], which was partially resolved with support from a non-governmental organization providing drivers and some financial and material resources.” (Study 2, Country 6, Trachoma, Lymphatic filariasis)*

*“…when dealing with security challenges in certain areas, we convene special planning meetings, involving traditional rulers, district representatives, LGAs, and experts from the water and sanitation sector…..We seek their insights and opinions, particularly regarding safe places to conduct our service interventions. If any areas are deemed unsafe, we take their advice seriously and adjust our plans accordingly. These planning meetings are invaluable as they complement our security checks and often provide crucial information that helps us navigate complex security situations effectively…. .It’s crucial for everyone to collaborate and contribute to ensuring safety during program implementation.” (Study 2, Country 7, trachoma)*

### External programme influences

There were five main external programme influence themes, which were all common to both studies: access to insecure areas, WASH, inclusivity, stakeholder partnering, and financial resources (Figure 1C, D and Table 2). There were six sub-themes; two were common between the studies (displaced populations and access to women), whereas Study 2 also raised the themes of impact of conflict, security, reliance on donors, and limited domestic resources (Figure 1). Selected key quotes are in Boxes 5-8.

### Access to insecure areas (Common theme across the two studies)

Ten participants expressed the barriers encountered by their NTD programmes when trying to gain access to areas of insecurity, whilst also maintaining the safety of their healthcare workers and programme officers. Participants highlighted the importance of collaborating with security personnel, to continually assess the security situation in their programme areas. Selected quotes in Box 5. These were also highlighted as potential solutions in Study 2 as highlighted in Box 4.

#### Impact of conflict (Study 2 sub-theme)

Six participants revealed the impact of conflict on NTD programme execution. They highlighted the difficulty of reaching conflict-affected areas due to unstable conditions, blockades, and personal security risks, which not only hamper activities but endanger their teams.

Displacement of large populations during conflicts exacerbates the spread of illnesses and obstructs disease management efforts. Additionally, achieving primary access within NTD programmes is complicated by cross-border issues and regional instability, leading to missed treatment cycles for affected populations.

#### Fieldwork Security (Study 2 sub-theme)

Three participants discussed the complications of involving security personnel for programme implementation in areas of conflict and relying on security forces for clearance to access various regions, adding complexity to their tasks. They highlighted that collaborating with military forces in unstable regions is challenging, requiring expertise and often generating apprehension among the local community.

###### Box 5. Selected quotes on access to insecure areas

*“We work in the most complex and security-challenged LGAs [Local Government Areas], requiring military approval for deploying resources.” Study 1, (Nigeria, Trachoma)*

*“So, before we do any activity in any security challenge area, we usually do a security assessment, to see if those areas, those districts are actually safe.” (Study 1, Nigeria, Trachoma)*

*“The teams now required the inclusion of a security personnel. Security considerations dictated our daily plans.” (Study 2, Country 3, Trachoma)*

***Impact of conflict***

*“Even currently there is an ongoing insecurity in different parts of the country. So, we have NTD care areas which are not currently accessible by the national program.” (Study 2, Country 1, all NTDs)*

*“Our primary challenges primarily revolve around cross-border issues, ongoing conflicts, and insecurities in the regions where we conduct our programs.” (Study 2, Country 5, Schistosomiasis, STH, Onchocerciasis lymphatic filariasis and trachoma)*

***Fieldwork security***

*“It led to increased costs due to the expanded team size. Previously consisting of only two individuals, the teams now required the inclusion of a security guard.” (Study 2, Country 4, trachoma, lymphatic filariasis, soil transmitted disease, snake bites, schistosomiasis, scabies)*

*“So, before we do any activity in any security challenged area, we usually do a security assessment, to see if those areas, those districts, are actually safe.” (Study 1, Nigeria, Trachoma)*

### WASH (Common theme across the two studies)

Twelve participants discussed the importance of sustaining and promoting WASH activities to support the outcomes of NTD programmes in conflict-affected areas, including working with WASH stakeholders at coordination level, to strengthen capacity for resources. This included the promotion of WASH related messages, to encourage hygiene practices and reduce rates of open defecation. Selected quotes in Box 6.

### Inclusivity (Common theme across the two studies)

Ten participants identified the importance of inclusivity for marginalised groups, such as internally displaced persons, refugees, and women, in the planning and implementation of their NTD programmes, particularly in areas of insecurity. Two participants highlighted the importance of leaving no one behind as they implement their initiatives. Selected quotes in Box 6.

#### Displaced populations (Common sub-theme)

Ten participants noted the difficulties identifying internally displaced populations and implementing successful NTD programmes in areas hosting IDPs, which are often areas of high poverty with unsanitary conditions due to their movement and not knowing how the exact population in the area.

#### Access for women (Common sub-theme)

Nine participants discussed the difficulties women face in accessing NTD treatment programmes, due to cultural norms of male doctors not treating female patients. Three participants noted benefit of recruiting female healthcare workers and surgeons to mitigate this. It was also noted that women face barriers accessing clean water, further exacerbating the spread of NTDs. Such issues were prevalent in areas of insecurity.

###### Box 6. Selected quotes on WASH and inclusivity

***WASH***

*“For us, the collaboration with security agencies, environmental ministry, and water resources ministry, we’ve been working well with them.” (Study 1, Nigeria, Trachoma)*

*“migration poses a significant challenge. Moreover, we grapple WASH, with low water and sanitation coverage.” (Study 2, Country 5, Trachoma, Guinea Worm, Mycetoma)*

***Inclusivity***

*“We do operate refugee camps in ……, hosting individuals from neighbouring war-torn countries such as, …., ……, and “These refugee camps are distinct entities when it comes to our Neglected Tropical Disease (NTD) programs.” (Study 2, Country 5, Trachoma, Guinea Worm, Mycetoma)*

*“Pinpoint the difficulties encountered, particularly in isolated communities where women may find it hard to obtain clean water, which can exacerbate the spread of neglected tropical diseases” (Study 2, Country 4, trachoma, lymphatic filariasis, soil transmitted disease, snake bites, schistosomiasis, scabies)*

*“In certain regions, we are strategically planning and collaborating with security forces to ensure that we can access hard-to-reach areas affected by conflicts, leaving no one behind as we implement our initiatives.” (Study 2, Country 5, Trachoma, Guinea worm, Mycetoma)*

***Displaced populations***

*“The goal is to help those who have fled from distressing situations, and it would be beneficial to understand the scale of the population affected by the conflict to plan interventions effectively. However, the issue is complicated by the fact that these displaced people do not remain stationary, which adds another layer of complexity to the situation.” (Study 1, Country 4, trachoma, lymphatic filariasis, soil transmitted disease, snake bites, schistosomiasis, scabies)*

### Stakeholder partnering (Common theme across the two studies)

Ten participants highlighted the significance of mapping and partnering with key stakeholders working in and around areas of insecurity to remove barriers to implementing successful NTD programmes. Key partnerships, including with the Ministry of Environment, Ministry of Education, and the Ministry of Water Resources, have greatly benefitted NTD programmes in areas of insecurity. Selected quotes in Box 7.

##### Box 7. Selected quotes on stakeholder partnering

***Stakeholder partnering***

*“We’ve been able to map some areas because we have to work with an International Organisation of Migration, and other such agents, UN agencies that help in you know, help to track people that are displaced.” (Study 1, Nigeria, Trachoma)*

*“We had cooperation with the security in the state, we had cooperation with the stakeholders, the state Ministries of Health, and various community leaders.” (Study 1, Nigeria, Trachoma)*

### Financial resources (Common theme across the two studies)

Seven participants highlighted the financial limitations NTD programmes face. This included the issue of organisation dependence on external funding because of limited domestic resources, particularly in countries experiencing ongoing conflicts or economic challenges and subsequent donors not investing their funding in the most effective areas of NTD programme. It was revealed that donors often fund MDA programmes but provide little funding for surveillance and impact assessment for programs in conflict-affected areas, which would have the greatest benefit. . Selected quotes in Box 8.

#### Reliance on donors (Study 2 sub-theme)

Four participants highlighted how NTD control programmes seek support from external funding sources like international organisations, NGOs, and bilateral donors. These entities provide financial backing for NTD control measures if a strategic plan and stability are in place. The interviewees highlighted their reliance on medication donations from pharmaceutical companies, especially for MDA. This dependence on external funding is often a result of limited domestic resources, particularly in countries experiencing ongoing conflicts or economic challenges.

#### Limited domestic resources (Study 2 sub-theme)

Two participants discussed that regions are often impacted by fluctuating economies, hindering governments from dedicating adequate resources to public health endeavours such as NTD control initiatives. These conflict areas often face competing financial priorities, which can lead to inadequate funding for healthcare, an issue that becomes especially pronounced in conflict-affected regions.

###### Box 8. Selected quotes on financial resources

***Financial resources***

*“Sustainability is now the issue. Because when these people are not there, I don’t know how some of the programme continues, even though we have made a lot of progress in terms of elimination.” (Study 1, Nigeria, Lymphatic filariasis, onchocerciasis, trachoma, schistosomiasis soil transmitted helminthiasis)*.

*“Our activities are also influenced by donor conditions, as some donors require stability before providing funds”. (Study 2, Country 4, trachoma, lymphatic filariasis, soil transmitted disease, snake bites, schistosomiasis, scabies)*

*“It led to increased costs due to the expanded team size. Previously consisting of only two individuals, the teams now required the inclusion of a security guard.” (Study 2, Country 4, trachoma, lymphatic filariasis, soil transmitted disease, snake bites, schistosomiasis, scabies)*

## Discussion

This paper sheds light on the substantial hurdles that NTD programmes face in conflict-torn areas of sub-Saharan Africa. The interviews with key informants were an effective qualitative approach for obtaining direct stakeholder information and their experiences on the additional pressures related to NTD control and elimination. We found a combination of common and unique challenges, which may be practically addressed through research and the development of a set of standardised strategies and sub-set of bespoke strategies to help meet the needs of individual countries. These may be considered together with other interconnected complex challenges and humanitarian emergencies [24].

The primary challenges specific to NTD programmes were found to be operational and logistical, related to activities such as surveillance, mapping, community education, and communication. The limited access to affected populations, damage to healthcare infrastructure, and poor communication technology impacted the delivery of essential services, as found with other diseases in other studies [7,25,26]. These were potentially exacerbated by population displacement, staff migration and shortages due to security risks. Selected programmes adapted to these circumstances, for example, by moving patients, or training residents and community health workers. The extent of the impact on health workers, their work, and highly vulnerable populations, such as refugees, is largely unknown and needs further research. The development of an NTD-specific framework, including risk assessments and mitigation action tools, is needed to help identify pathways of threats, and methods of adaptation, community engagement, and programme and health worker optimisation. Lessons may be learned from other programmes [27–30].

Security risks are the most pressing external challenge, with unpredictable violence and blockades restricting access to endemic areas, which increases the risk of programmes being unable to ‘leave no one behind’ and meet global NTD goals [13,30–32]. Conflict-related displacement complicates the situation as populations are unstable, and often move to areas that lack basic resources and services, such as WASH. This can increase the risk transmission and of other infectious diseases such as cholera, which may divert human and financial resources [6,9,33,34].

Recent evidence in Nigeria has examined ways to improve inclusiveness and access for displaced populations, highlighting success through collaboration with a State Emergency Management Agency to map camps, and the training of displaced persons to improve access to treatment [30].

The dependency on external funding leaves NTD and other global health programmes vulnerable to shifts in international financial support exemplified in dramatic reductions in US funding to WHO [35]. These external pressures necessitate urgent attention to health financing, resource mobilization, and cross-sectoral collaboration, which align with the three fundamental pillars of the NTD Road Map 2021-2030; i) accelerating programmatic action ii) intensifying cross-cutting approaches; and iii) changing operating models and culture to facilitate country ownership [2]. Being aware of how donor priorities may change is important, and forging new collaborations with the WASH and humanitarian sectors to maximise limited resources warrants further consideration [24]. The critical linkage between NTDs and WASH has already been recognised, with a global strategy, guidelines, and other key updates available[36]. These need to be promoted in conflict areas.

Addressing the challenges in conflict zones requires a combination of strategic interventions. Conflict resolution efforts, such as peace-building initiatives, are essential to ensure the safety of healthcare staff and accessibility to affected regions [37]. These are out of the scope of NTD programmes; however, their efforts can help to strengthen health worker preparedness, and healthcare infrastructure, including logistics management and communication systems. In addition, collaboration with the education sector and the training and adoption of new technologies and innovations will support healthcare workforce [24,38,39]. There are a wide variety of resources available on the WHO learning resource hub (https://openwho.org/) on emergencies, including preparedness and response framework, and capacity strengthening as a starting point for crisis management.

Moreover, fostering stronger partnerships with stakeholders, including UN agencies and NGOs from a variety of sectors, is vital to creating sustainable solutions for NTD control [24]. It was reported in this study that NTD programmes in Nigeria partnered with security operators to conduct security assessments, to determine when and how these areas can be entered. Collaboration with security personnel have been shown to be effective for other health programmes such as in Borno State of Nigeria where it increased access to insecure areas and improved evaluation indicators for polio programmes [26].. As such, participants in this study highlighted how vital it is to partner with security personnel working in areas of insecurity, to enable the safest and most effective interventions.

The main limitations to this study included its potential biases arising from language barriers, relatively small sample size, and the focus on countries with high conflict levels, which may limit the generalisability of the findings. Notwithstanding these limitations, use of purposive sampling and digital online interviews offered a flexible, cost-effective foundation for further studies to be conducted in similar contexts across many countries. Further, the study will help develop policies and guidelines related to mapping, surveillance, and response strategies in conflict-affected areas [11,12], and identify key research priority themes, such as those summarised in Table 3, related to resilience, knowledge and capacity, vulnerable populations, new technologies and community.

**Table 3.** Summary of key research priority areas in conflict-affected areas.

| Priority area | Description |
| --- | --- |
| NTD specific framework | Develop a practical framework by determining what strategies are needed, who needs to be involved, and how they may be implemented to enable programmes to better mitigate, prepare, respond and recover and report. |
| Knowledge transfer and capacity strengthening | Develop effective materials and methods for educating and disseminating best practices and building local expertise and capacity that are bespoke to unique challenges. |
| Vulnerable populations and equity | Research on the healthcare and programme needs of vulnerable populations, including displaced and marginalised communities, to address inequalities. |
| New technologies and innovation | Assess the potential and scale up of new technologies, such as digital tools, to support diagnosis, treatment, reporting, management, and communication. |
| Community engagement | Evaluate the effectiveness of community-centred solutions, and local health representation and participation for improving NTD programme activities. |

## Conclusion

NTD programmes in conflict-affected sub-Saharan African countries face many interconnected programme-specific and external challenges. Key solutions include cross-sectoral partnerships, community engagement, health system strengthening, innovation and research. There is an urgent need to develop practical strategies to support programmes so that progress can be made towards the NTD road map 2021-2030 control and elimination goals.

## Data Availability

All data produced in the present work are contained in the manuscript.

## Authors’ contributions

LAKH and EMHE conceived the study and contributed to the study design. FB (study 1) and AB (study 2) designed the study protocol and collected and analysed the data. FB, AB, LAKH and EMHE interpreted the data and drafted the manuscript. All authors read and approved the final version.

## Acknowledgements.

We thank members of the NTD NGO Network Conflict and Humanitarian Emergency working group and International Coalition for Trachoma Control task team for reviewing the topic guides.

## Ethical approval

Ethical approval was obtained through the London School of Hygiene & Tropical Medicine (Ethics Ref: 28621 and 29060) and the University of Liverpool Ethics committees (Ethics Ref: 12760). Local approval was received from the Nigerian Federal Ministry of Health through a letter of support.

## Supplementary file

### Study 1 - Semi-Structured Interview Topic Guide

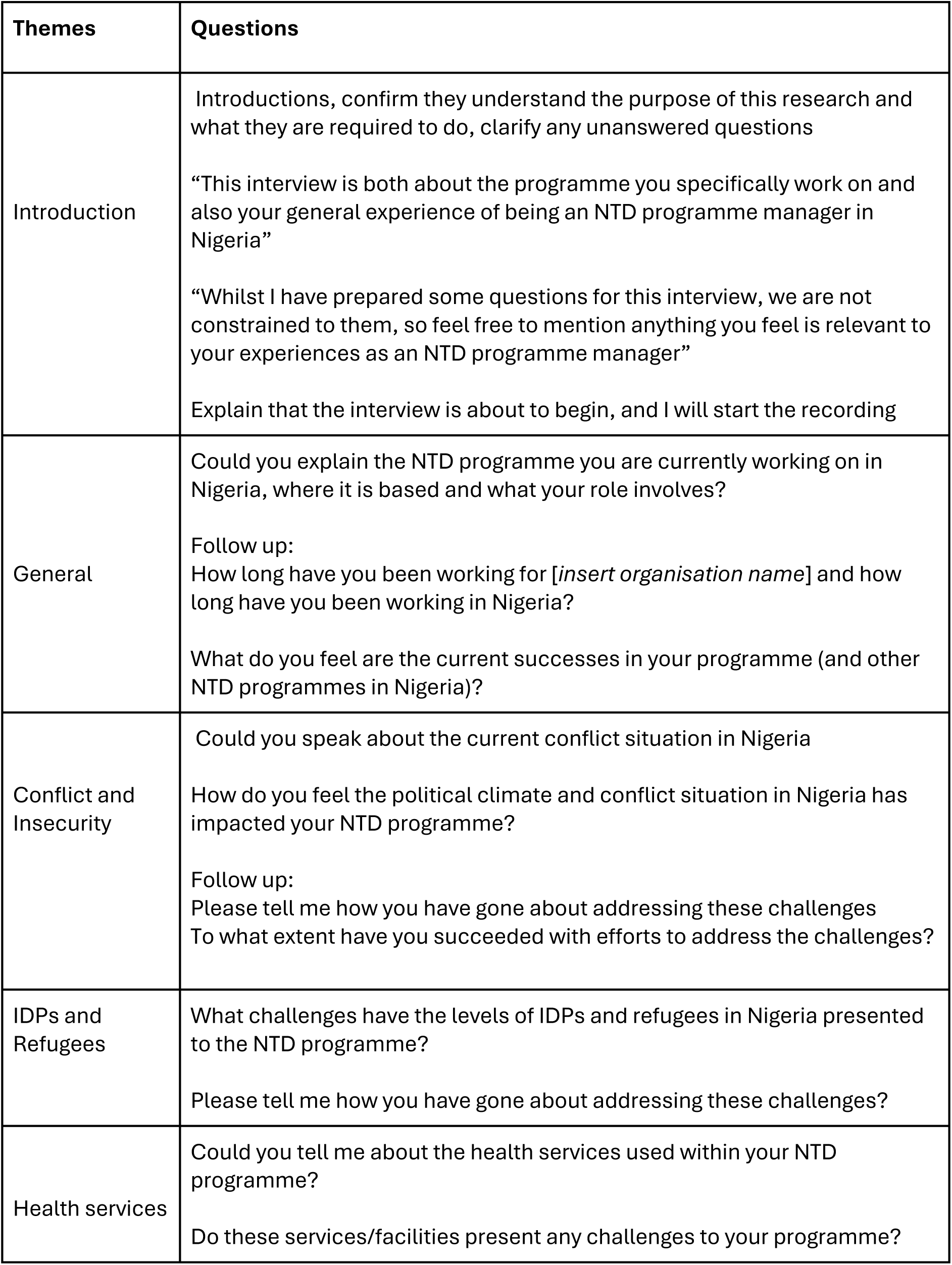
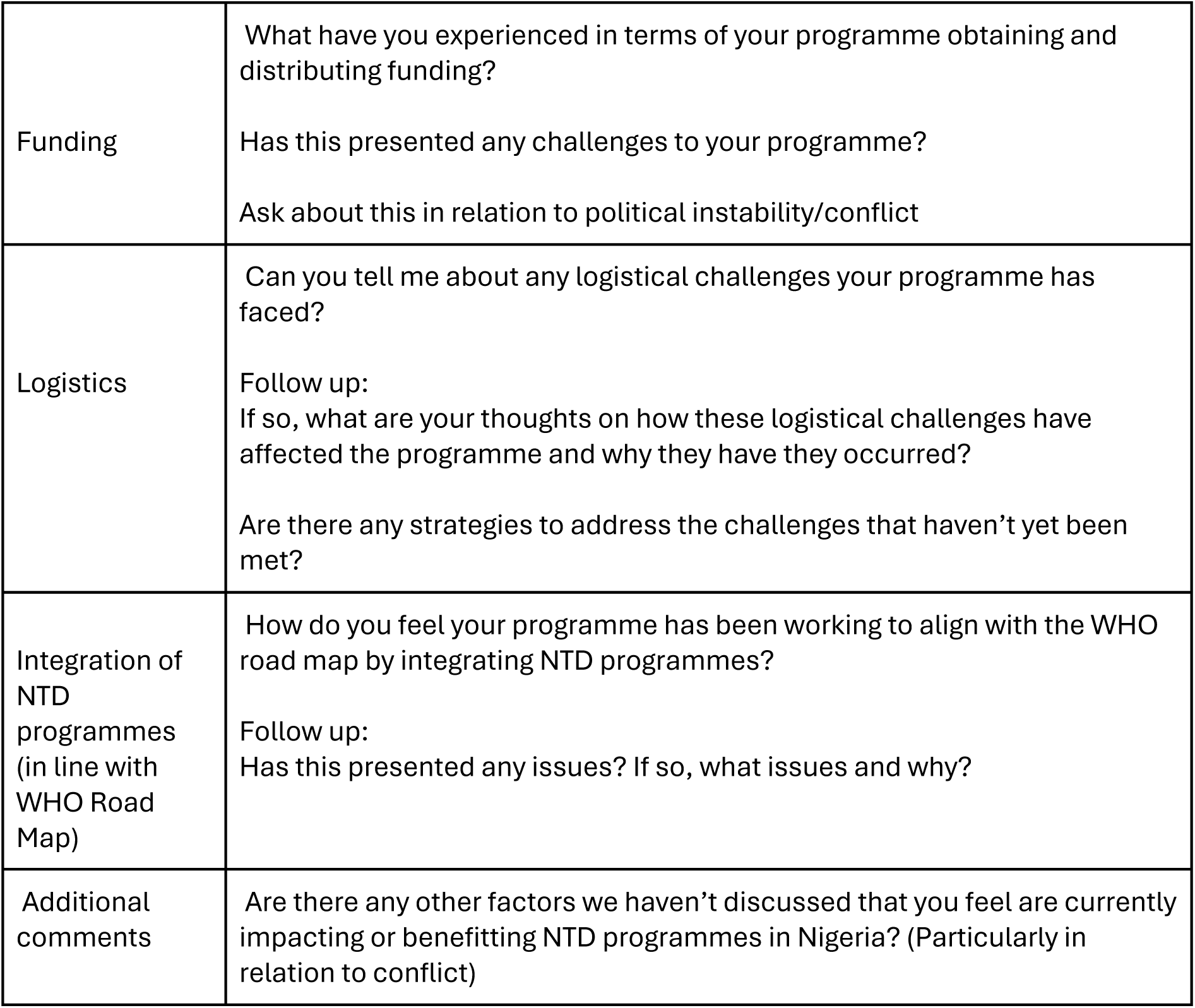

| Themes | Questions |
| --- | --- |
| Introduction | Introductions, confirm they understand the purpose of this research and what they are required to do, clarify any unanswered questions<br>“This interview is both about the programme you specifically work on and also your general experience of being an NTD programme manager in Nigeria”<br>“Whilst I have prepared some questions for this interview, we are not constrained to them, so feel free to mention anything you feel is relevant to your experiences as an NTD programme manager”<br>Explain that the interview is about to begin, and I will start the recording |
| General | Could you explain the NTD programme you are currently working on in Nigeria, where it is based and what your role involves?<br>Follow up:<br>How long have you been working for [*insert organisation name*] and how long have you been working in Nigeria?<br>What do you feel are the current successes in your programme (and other NTD programmes in Nigeria)? |
| Conflict and Insecurity | Could you speak about the current conflict situation in Nigeria<br>How do you feel the political climate and conflict situation in Nigeria has impacted your NTD programme?<br>Follow up:<br>Please tell me how you have gone about addressing these challenges<br>To what extent have you succeeded with efforts to address the challenges? |
| IDPs and Refugees | What challenges have the levels of IDPs and refugees in Nigeria presented to the NTD programme?<br>Please tell me how you have gone about addressing these challenges? |
| Health services | Could you tell me about the health services used within your NTD programme?<br>Do these services/facilities present any challenges to your programme? |
| Funding | What have you experienced in terms of your programme obtaining and distributing funding?<br>Has this presented any challenges to your programme?<br>Ask about this in relation to political instability/conflict |
| Logistics | Can you tell me about any logistical challenges your programme has faced?<br>Follow up:<br>If so, what are your thoughts on how these logistical challenges have affected the programme and why they have they occurred?<br>Are there any strategies to address the challenges that haven't yet been met? |
| Integration of NTD programmes (in line with WHO Road Map) | How do you feel your programme has been working to align with the WHO road map by integrating NTD programmes?<br>Follow up:<br>Has this presented any issues? If so, what issues and why? |
| Additional comments | Are there any other factors we haven't discussed that you feel are currently impacting or benefitting NTD programmes in Nigeria? (Particularly in relation to conflict) |

### Study 2 - Semi-Structured Interview Topic Guide

#### Topic Guide

*Introduction:*

*Introduction and explain the purpose of the study.*

##### Section 1: Information about participant

1. How long have you been working as an NTD program manager in a conflict-affected African country?
2. Which NTDs do you work on?
3. What does your role involve?

##### Section 2 NTD control programs in conflict-affected countries

1. Can you describe the NTD control program in your country and how it is implemented?
2. What are the main challenges you face in implementing the NTD control program in a conflict-affected setting?

##### Section 3 Overcoming Challenges

1. Can you describe the main challenges you face while implementing the NTD control program in a conflict-affected setting?
2. How do you overcome these challenges?
3. Were there any unexpected outcomes or benefits that arose because of addressing these challenges?

##### Section 4 Potential solutions

1. In your opinion, what are some potential solutions that could improve the effectiveness of NTD control programs in conflict-affected African countries?
2. Have you implemented any innovative or effective solutions in your own NTD control program?
3. What resources or support would be necessary to implement these solutions? (Probe on types of resources, which organisations they would need support from)

#### Conclusion

1. *Thank the participant for their time and insights*.
2. *Confirm that the recording will be kept confidential and anonymized*.
3. *Inform the participant that they can withdraw their consent or request to have their data removed from the study at any time*.

